# Metabolomic profiling in treated mucopolysaccharidosis IH reveals candidate biomarkers and adjunctive therapeutic pathways

**DOI:** 10.64898/2026.09.04.26362233

**Authors:** Troy C. Lund, Ryan H. Peretz, Patricia I. Dickson, Jennifer K. Yee, Michelina Iacovino, Kent D. Taylor, David Elashoff, Ellen Fung, Bradley S. Miller, Paul J. Orchard, Lynda E. Polgreen

## Abstract

Both severe (Hurler syndrome; MPS IH) and attenuated (Hurler-Scheie or Scheie syndrome; MPS IA) forms of mucopolysaccharidosis I (MPS I) arise from the same underlying enzyme deficiency; however, they differ in the onset, persistence, and severity of key clinical features. These include skeletal abnormalities, joint contractures, cardiac disease, and neurocognitive and neurobehavioral impairment, which is not fully alleviated with either enzyme replacement (ERT) or hematopoietic cell transplantation (HCT). The objective of this study was to identify metabolic differences between MPS IH and MPS IA that could result in meaningful biomarkers and targeted adjunctive therapies to address unmet clinical needs in MPS IH treated with HCT. We performed plasma metabolomics in patients with MPS IH treated with HCT (N=17) or MPS IA treated with ERT (N=8). Welch’s two-sample t-test was used to identify metabolites that differed significantly between groups. Statistical significance was evaluated based on p<0.05. After controlling for multiple testing, we used a false discovery rate of q<0.05. We identified 125 compounds that were significantly different between MPS IH and MPS IA. Of those metabolites, 14 had a q<0.05. Individuals with MPS IH had increased metabolites in the sphingolipid, beta-oxidation, amino acid catabolism, and glycosaminoglycan pathways compared to MPS IA. Persistent metabolic differences were observed in MPS IH treated with HCT compared to MPS IA treated with ERT, pointing towards additional biological processes that may contribute to disease progression in MPS IH after HCT. Although these results require further confirmation, they provide a foundation to guide future investigations towards potential biomarkers or targeted adjunctive therapies.

## 1. INTRODUCTION

Mucopolysaccharidosis type I (MPS I) is a lysosomal storage disease caused by variants in the *IDUA* gene that manifest as central nervous system (cognitive impairment) and peripheral abnormalities (bone, joint, heart, liver) that vary broadly in severity and symptom onset[1]. Patients with MPS I can have relatively mild clinical manifestations and live reasonably normal lifespans with near normal cognition, or they can have a rapidly progressing fatal course within the first two decades of life. The severe form is called MPS IH (Hurler syndrome; OMIM 607014), and the attenuated form is MPS IA (encompassing both Hurler-Scheie, OMIM 607015; and Scheie, OMIM 607016, syndromes).

These distinct forms of MPS I are treated differently. Children with MPS IH require hematopoietic cell transplantation (HCT), often with peri-HCT enzyme replacement therapy (ERT), because HCT can alter the course of neurologic deterioration when performed in children prior to the onset of neurocognitive disability[2–4]. The increased risks associated with HCT are typically thought not justified for individuals with MPS IA due to the relative lack of neurodegenerative disease, thus the disorder is managed with ERT. These treatments are not cures. Most individuals with MPS I continue to live with varying degrees of CNS disease, cardiac valvular disease, chronic pain and limitations in their activities of daily living due to persistent joint dysfunction and skeletal dysplasia[2–15].

The discovery of downstream effects of lysosomal dysfunction resulting from the accumulation of abnormal glycosaminoglycans (GAGs) in MPS has inspired targeted adjunctive therapies for persistent disease manifestations. For example, a recent transcriptomic analysis of fibroblasts from patients with neuronopathic MPS, compared to non-neuronopathic MPS and healthy controls, identified a gene expression pattern in neuronopathic MPS suggesting abnormalities in the structure and function of the cell nucleus, endoplasmic reticulum, and Golgi apparatus[16]. Additionally, preclinical studies have shown that incompletely degraded GAGs released into the extracellular matrix can activate innate immune responses through interleukin-1 (IL-1)[17–19] and toll-like receptor 4 (TLR4)[20–22] signaling. This leads to neuroinflammation and synovial changes that mirror those seen in neurologic autoinflammatory and inflammatory joint diseases. Therapeutically targeting these pathways has yielded promising results in animal models, including reductions in neuroinflammation, improvements in cognitive and behavioral symptoms[19], and decreased joint inflammation with improved mobility[21,22]. These mechanistic insights provided the foundation for clinical trials of adjunctive therapies. For example, anti-TNF-α treatment in individuals with MPS I and II resulted in improved joint mobility and reduced chronic pain[23], while anti-IL-1 receptor antagonist therapy in MPS III led to improvements in behavioral and functional outcomes[24]. Together, these findings highlight the importance of identifying persistent physiological abnormalities in mucopolysaccharidoses as a critical step toward expanding adjunctive, pathway-specific therapeutic options. Thus, the aim of this study was to pursue metabolomic profiling to evaluate differences between MPS IH treated with HCT and MPS IA treated with ERT to generate hypotheses of potential biomarkers and targeted adjunctive treatments for the disease manifestations resistant to HCT in MPS IH.

## 2. MATERIALS AND METHODS

### 2.1 Study Population

Written informed consent was obtained from all parents/guardians of the subjects and assent from subjects cognitively able to provide assent. Institutional Review Boards at all study sites approved the protocol. Plasma samples from 25 participants with MPS I (17 with MPS IH; 8 with MPS IA), age 5.0 to 20.8 years, were enrolled in this 10-year, multi-center, longitudinal observational study (**Table 1**). Samples were collected from September 24, 2009 to May 1, 2014. Despite the significant difference in age between the two groups, there was no significant difference in BMI percentile, which would have impacted metabolomic profiles in adolescents [25]. One participant with MPS IH had not previously undergone HCT; all others had been treated with HCT more than two years prior to metabolome profiling. All participants with MPS IA were being treated with ERT for more than one year at study initiation. Inclusion criteria were diagnosis of MPS I, age 5 to 33 years of age, and ability to travel to the study center. Exclusion criteria were pregnancy, participation in any other study within the past 6 months that would increase radiation exposure above 500 mrem for the calendar year, and inability to comply with study procedures. Venous samples were collected in 4 mL EDTA tubes in the morning, after a minimum 8-hour fast, and immediately processed and stored at a central institution at -80°C. Samples were thawed, aliquoted, and immediately restored at -80°C prior to sending to Metabolon Incorporated for metabolomics analysis.

**Table 1.** Demographic characteristics of study cohorts.

| **Table 1. Demographic characteristics of study cohorts.** |  |  |
| --- | --- | --- |
|  | **MPS IH**<br>**N=17** | **MPS IA**<br>**N=8** |
| Age – years |  |  |
| Mean ± SD | 9.5 ± 3.3 | 15.2 ± 3.5 |
| Median | 10.6 | 15.8 |
| Range | 5.0 - 14.5 | 8.9 - 20.8 |
| Treated with HCT – no. (%) | 16 (94) | 0 (0) |
| Age at HCT initiation – years |  |  |
| Mean ± SD | 1.3 ± 0.7 | - |
| Median | 1.2 |  |
| Range | 0.2 - 2.9 |  |
| Treated with ERT – no. (%) | 7 (41) | 8 (100) |
| Age at ERT initiation – years |  |  |
| Mean ± SD | 1.3 ± 0.7 | 8.9 ± 2.6 |
| Median | 1.2 | 8.8 |
| Range | 0.5 - 2.4 | 5.7 - 13.0 |
| BMI – percentile |  |  |
| Mean ± SD | 66.5 ± 25.8 | 53.5 ± 13.0 |
| Median | 63.1 | 58.1 |
| Range | 16.9 – 97.0 | 2.7 – 99.6 |
| Time since HCT – years |  |  |
| Mean ± SD | 8.4 ± 3.2 | - |
| Median | 8.8 | - |
| Range | 3.9 - 12.5 | - |
| Time since ERT initiation – years |  |  |
| Mean ± SD | - | 6.3 ± 3.8 |
| Median | - | 5.5 |
| Range | - | 1.9 - 11.9 |
| Sex, female – no. (%)* | 9 (53) | 3 (38) |
| Race – no. (%)* |  |  |
| White | 17 (100) | 6 (75) |
| Black | 0 (0) | 1 (12.5) |
| American Indian | 0 (0) | 1 (12.5) |
| Ethnicity – no. (%)* |  |  |
| Hispanic | 2 (12) | 1 (13) |
| Non-Hispanic | 15 (88) | 7 (87) |
| *Sex, Race, and Ethnicity were reported by the participant and/or participant's parent/caregiver.<br>MPS IH = Hurler syndrome (severe MPS I); MPS IA = Hurler-Scheie and Scheie syndrome (attenuated MPS I); HCT=hematopoietic cell transplantation; ERT=enzyme replacement therapy |  |  |

### 2.2 Metabolomics

Untargeted metabolomic analysis was performed at Metabolon Incorporated (Durham, NC) via standardized procedures and quality control measures (**S1 Supplemental Methods**). Briefly, raw data was extracted, peak-identified, and quality control-processed at Metabolon using Metabolon’s hardware and software. More than 3300 commercially available purified standard compounds have been acquired and registered into the Laboratory Information Management System (LIMS) for analysis on all platforms for determination of their analytical characteristics. Additional mass spectral entries have been created for structurally unnamed biochemicals, which have been identified by their recurrent nature (both chromatographic and mass spectral). Metabolon data analysts use proprietary visualization and interpretation software to confirm the consistency of peak identification among the various samples. Peaks were quantified using area-under-the-curve.

### 2.3 Statistical Analysis

Values were median scaled, using the median for the run day of each sample, on a per metabolite basis. Those median scaled values were then log transformed prior to statistical analysis. Any sample that did not have a peak for a given metabolite was imputed with the sample set minimum value. Welch’s two-sample t-test was used to identify metabolites that differed significantly between experimental groups. An estimate of the false discovery rate (q-value) was calculated to account for the multiple comparisons that normally occur in metabolomic-based studies (“Array Studio” R package)[26]. Initial significance was evaluated based on p<0.05. After controlling for multiple testing, we used a false discovery rate of q<0.05. We used the random forest method to compute the variable importance measure, and we rank ordered each compound based on this measure[27]. Data are presented in figures as a fold change representing the ratio of the average of the median scaled metabolite value in MPS IH relative to MPS IA.

## 3. RESULTS

Of 687 metabolites tested, the top 30 most important metabolites identified by random forest analysis are shown in **Fig 1**. Of these, four metabolites, namely methionine sulfone, 1-palmitoyl- 2-linoleoyl-GPI (16:0/18:2), N-acetylhistidine, and 1-methylimidazoleacetate best discriminated between participants with either MPS IH or MPS IA. The amino acid and lipid metabolic pathways were overall most involved in separation of groups.

**Fig 1.**
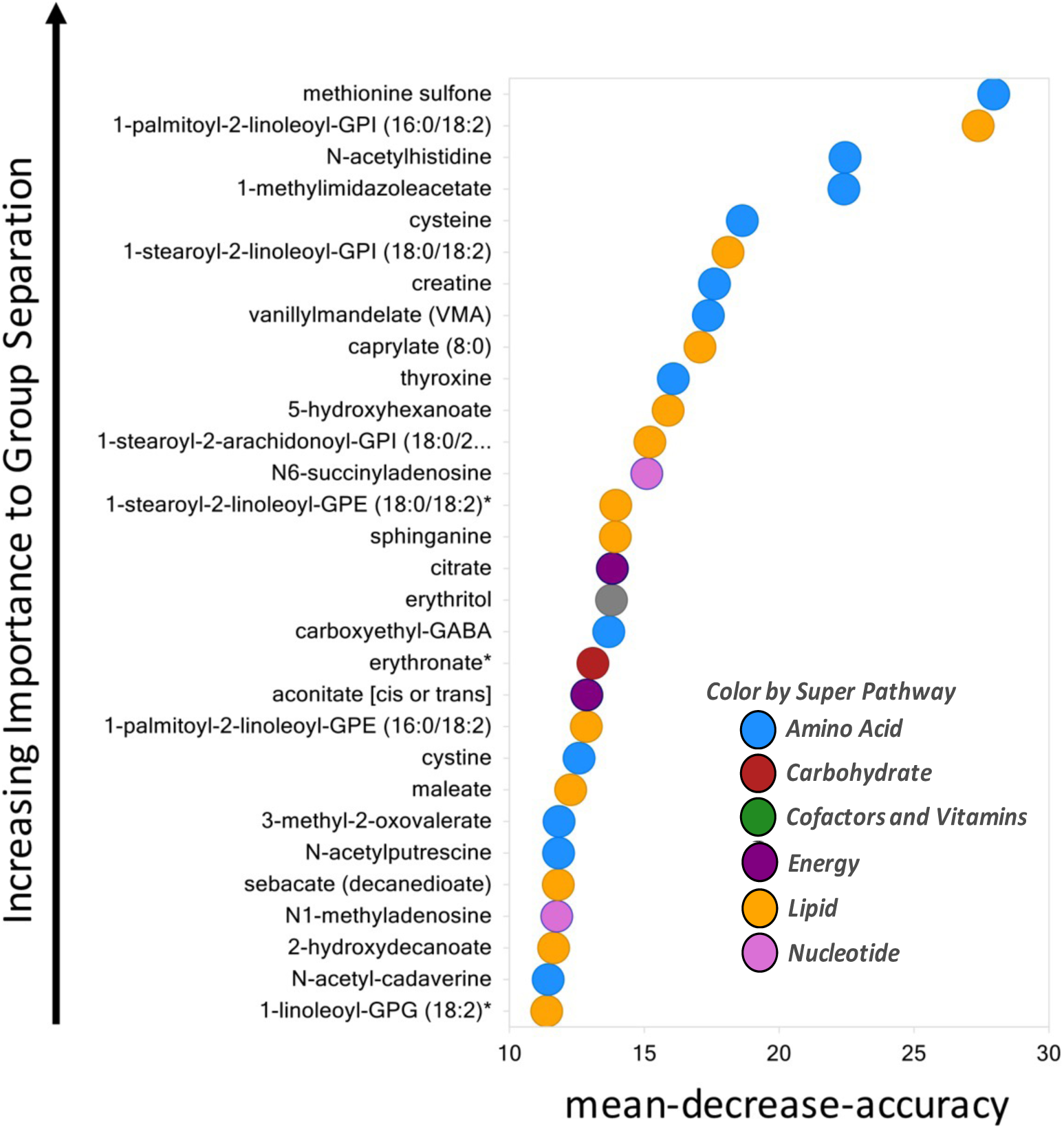
Random forest classification of metabolites that best distinguish between MPS IH and MPS IA. GPI = glycosylphosphatidylinositol; GABA = gamma-amino butyric acid; GPE = glycerophospholipid; GPG = glycerophosphoglycerol. * indicates compounds that have not been officially confirmed based on a standard, but Metabolon is confident in its identity.

The comparative metabolomic analysis between MPS IH and MPS IA revealed distinct metabolic perturbations. We identified 125 compounds that were significantly different (p<0.05; Welch’s two-sample t-test) between MPS IH and MPS IA (**S2 Supplemental Table)**; of those candidate metabolites, 14 also had a q<0.05 (**Table 2**). These 125 metabolites suggest dysregulation across several major biochemical pathways. Most notably, multiple sphingolipid pathway metabolites were increased in MPS IH. Metabolites involved with β-oxidation (including dicarboxylic acids, acyl-carnitines, hydroxy fatty acids), various lipid intermediates (phospholipids, lysolipids, monoacylglycerols, diacylglycerols), and urea cycle and polyamine metabolites (creatine, homocitrulline, 4-acetamidobutanoate) were significantly elevated in MPS IH. Amino acid catabolism also differed substantially between the two forms of MPS I. In MPS IH, elevated metabolites were observed across several amino acid metabolic pathways, including glutamate, histidine, lysine, phenylalanine, tyrosine, tryptophan, methionine, and cysteine.

**Table 2.** Metabolites with a q-value < 0.05 in comparative metabolomic analysis.

|  |  |  |  |  |  |  |
| --- | --- | --- | --- | --- | --- | --- |
| **Biochemical Name** | **KEGG** | **HMDB** | **PUBCHEM** | **Fold Change MPS IH/ MPS IA** | **p-value** | **q-value** |
| 1-palmitoyl-2-linoleoyl-GPI (16:0/18:2) |  |  |  | 1.98 | 7.83E-07 | 0.0002 |
| 1-stearoyl-2-linoleoyl-GPI (18:0/18:2) |  |  |  | 1.78 | 6.29E-05 | 0.0056 |
| cysteine | C00097 | HMDB00574 | 5862 | 1.73 | 6.03E-05 | 0.0056 |
| N-acetylhistidine | C02997 | HMDB32055 | 75619 | 2.08 | 0.0001 | 0.0096 |
| 1-methylimidazoleacetate | C05828 | HMDB02820 | 75810 | 1.89 | 0.0003 | 0.0112 |
| vanillylmandelate (VMA) | C05584 | HMDB00291 | 1245 | 1.8 | 0.0002 | 0.0112 |
| 5-hydroxyhexanoate |  | HMDB00525 | 170748 | 2.41 | 0.0004 | 0.0139 |
| carboxyethyl-GABA |  | HMDB02201 | 2572 | 2.02 | 0.0008 | 0.0251 |
| methionine sulfone |  |  | 69961 | 2.27 | 0.0008 | 0.0252 |
| N-acetylputrescine | C02714 | HMDB02064 | 122356 | 1.66 | 0.0012 | 0.0300 |
| 1-stearoyl-2-arachidonoyl-GPI (18:0/20:4) |  |  |  | 1.55 | 0.0012 | 0.0300 |
| pyridoxate | C00847 | HMDB00017 | 6723 | 1.96 | 0.0019 | 0.0423 |
| erythronate* |  | HMDB00613 | 2781043 | 1.34 | 0.0021 | 0.0423 |
| 3-aminoisobutyrate | C05145 | HMDB03911 | 64956 | 1.98 | 0.0026 | 0.0488 |
| GPI = glycosylphosphatidylinositol; KEGG = Kyoto Encyclopedia of Genes and Genomes; HMDB = Human Metabolome Database; GABA = Gamma-Aminobutyric Acid<br>* = Indicates compounds that have not been officially confirmed based on a standard, but Metabolon is confident in its identity. |  |  |  |  |  |  |

## 4. DISCUSSION

Here, we identified distinct metabolic alterations—most notably increased lipid and amino acid metabolic intermediates—in severe (MPS IH) compared to attenuated (MPS IA) MPS I among treated individuals. These findings reveal systemic metabolic differences that offer new insights into the biological consequences of lysosomal enzyme deficiency. The observed shifts in energy metabolism, amino acid catabolism, and lipid homeostasis may suggest broad ongoing cellular stress responses and compensatory adaptations to ongoing lysosomal dysfunction despite treatment with HCT.

Prior studies have evaluated metabolites in blood and/or urine of patients with MPS II and MPS III compared with healthy controls and, similar to our results in MPS IH, found alterations in amino acid metabolism [28–30]. Several groups have observed alterations in amino acid levels - arginine, aspartic acid, glutamic acid, tyrosine, and phenylalanine - in MPS III patients [26,27].

In the case of aspartic acid, one group found aspartic acid was markedly elevated in MPS III patient urine [29], whereas another group reported that it was low in serum samples from MPS III patients [28]. In MPS II patients, short peptides such as aspartate-arginine-arginine-tyrosine in urine and phenylalanylglycine in plasma suggest incomplete protein digestion in affected individuals compared with controls [30].

Studies have also identified elevated biomarkers of oxidative stress in MPS, and specifically disruptions in methionine metabolism [31,32]. In our analysis, methionine sulfone—the oxidized derivative of methionine—ranked highest in the Random Forest model for separating MPS IH from MPS IA. Methionine sulfone is produced through the oxidation of methionine residues and can be reversed by the enzyme methionine sulfoxide reductase, a key component of the anti- oxidant defense system[33]. Elevated methionine sulfone has been associated with accelerated disease progression in patients with other types of MPS (i.e., MPS II and MPS IV)[34–36] and has also been implicated in aging and other neurodegenerative diseases[37–40] suggesting a role for it in the development of more severe neurologic disease in MPS IH versus MPS IA. This interpretation is supported by prior findings in an MPS I mouse model (*Idua-/-* mice), which have upregulation of antioxidant enzymes such as catalase and superoxide dismutase[41], and by reports of increased oxidative stress biomarkers in untreated MPS I (subtype not specified)[31] and ERT-treated MPS I (predominantly MPS IA)[42] compared to healthy controls. Importantly, our data indicate that oxidative damage remains a key driver of disease burden even after HCT therapy for MPS IH.

Elevations in glycosylphosphatidylinositol (GPI)-tagged phospholipids were identified in our Random Forest analysis. Specifically, increased levels of 1-palmitoyl-2-linoleoyl-GPI (16:0/18:2), 1-stearoyl-2-linoleoyl-GPI (18:0/18:2), and 1-stearoyl-2-arachidonoyl-GPI (18:0/20:4) suggest an accumulation of GPI-anchor biosynthetic intermediates in the plasma. This likely reflects impaired processing or trafficking of GPI-anchored molecules such as glypicans 1-4 due in part to the inability to remove heparan sulfate from these GPI-anchors or the inability of other GPI anchored proteins to interact with heparan sulfates in the extracellular matrix[43–45]. GPI-anchors are critical for tethering proteins to membrane microdomains such as lipid rafts and are trafficked through the endosomal-lysosomal system [46–48]. Disruption of these processes could impair membrane organization, protein localization, and signaling, especially in the immune response. Secondary lipid accumulation is well described in MPS and other lysosomal storage diseases[49–51]. Similar phospholipid abnormalities have been observed in neuronopathic MPS II[52], supporting the relevance of these findings.

We found increased 1-methylimidazoleacetate and N-acetylhistidine in MPS IH compared to MPS IA, which are potentially relevant to histamine metabolism. Histamine is produced in humans by decarboxylation of histadine by histadine decarboxylase. Mast cells, which produce and contain histamine, were previously reported to contain ‘worm-like’ inclusions in skin biopsies from 6 children with MPS IH or MPS II[53]. In mammalian brains, histamine is converted to telemethyl-histamine via histamine-*N*-methyltransferase and is then metabolized into 1-methylimidazoleacetate via monoamine oxidase-B subtype[54]. However, elevations in our MPS IH cohort of N-acetylhistidine are of unclear etiology. Past research has established this molecule as being present in the human lens but without a known role in the human brain [55–57]. N-acetylhistidine is metabolized via histidine N-acetyltransferase, an enzyme that has only been observed in fish and reptiles. A potential human ortholog is *NAT16*, which codes for N-acetyltransferase 16[58]. In fish, there are notably high concentrations of histidine and histidine-related compounds in the brain to help with cerebral osmoregulation, pH buffering, and possible neuroprotective effects including antioxidant activity and stabilizing cell structures[59]. There have also been reports of N-acetylhistidine helping prevent cataracts in fish through its role in osmoregulation and serving as a storage form of histidine[55,59]. More research is clearly needed to clarify the role of circulating N-acetylhistidine in human biology and MPS IH specifically.

Study limitations include a small sample size that limits statistical power and the generalizability of the findings (e.g., all participants were White) and confounding that could occur with age differences between MPS IH and MPS IA groups. As a cross-sectional analysis, the study does not capture temporal changes or allow for causal inferences. The lack of integrated multi-omics data, including transcriptomics or proteomics, makes it difficult to determine whether these effects result from reduced catabolic capacity or enhanced anabolic activity. We are also unable to state with certainty that the metabolic alterations in MPS IH are due to MPS or a result from HCT; although since the standard of care for MPS IH is currently HCT at as young of an age as possible, this limitation has limited clinical importance. It is also unknown whether this study’s findings represent pathophysiology, or adaptive metabolic shifts in response to the pathophysiology.

## CONCLUSIONS

In summary, our study revealed distinct and measurable metabolic differences—particularly in lipid and amino acid metabolism—between treated individuals with MPS IH and MPS IA. Persistent, phenotype-specific metabolic differences, despite treatment, points to underlying biological processes that may contribute to differences in disease progression. Although these results require further confirmation and investigations to draw definitive conclusions, they may help inform future directions in the development of biomarkers or adjunctive therapies in MPS IH after HCT.

## Data Availability

All data generated or analyzed during this study are included in this published article and its supplementary information files.

## ACKNOWLEDGMENTS

This project was supported by a grant from Genzyme/Sanofi (LEP), and grant numbers K23AR057789 (LEP) from the National Institute of Arthritis and Musculoskeletal and Skin Diseases (NIAMS), U54NS065768 (LEP) from the National Center for Advancing Translational Sciences (NCATS), the National Institute of Neurological Disorders and Stroke (NINDS), and the National Institute of Diabetes and Digestive and Kidney Diseases (NIDDK), and by UL1TR000124, UL1TR000114 and UL1RR024131 from NCATS of the National Institutes of Health (NIH) to the University of California – Los Angeles, the University of Minnesota and the Children’s Hospital Oakland Research Institute, respectively, Clinical and Translational Science Institutes (CTSI). The Lysosomal Disease Network (U54NS065768) is a part of the Rare Diseases Clinical Research Network (RDCRN), an initiative of the Office of Rare Diseases Research (ORDR), and NCATS. This consortium is funded through a collaboration between NCATS, NINDS, and NIDDK. Its contents are solely the responsibility of the authors and do not necessarily represent the official views of the CTSI or the NIH.

## AUTHOR CONTRIBUTIONS

Ryan H. Peretz: Writing – Review & editing, Visualization

Troy C. Lund: Conceptualization, Methodology, Resources, Writing – Review & Editing Patricia I. Dickson: Writing – Review & Editing

Jennifer K. Yee: Writing – Review & Editing Michelina Iacovino: Writing – Review & Editing Kent D. Taylor: Writing – Review & Editing David Elashoff: Writing – Review & Editing

Ellen Fung: Investigation, Resources, Writing – Review & Editing, Supervision, Project Administration

Bradley S. Miller: Investigation, Resources, Writing – Review & Editing, Supervision, Project Administration

Paul J. Orchard: Methodology, Resources, Writing – Review & Editing

Lynda E. Polgreen: Conceptualization, Methodology, Investigation, Resources, Data Curation, Writing – Original Draft, Visualization, Supervision, Project Administration, Funding Acquisition

## DECLARATION OF GENERATIVE AI IN SCIENTIFIC WRITING

During the preparation of this work the author (LEP) used ChatGPT in order to improve readability. After using this tool/service, all authors reviewed and edited the content as needed and take full responsibility for the content of the publication.

## S1 Supplemental Methods

### Metabolon Platform

#### Sample Accessioning

Following receipt, samples were inventoried and immediately stored at -80°C. Each sample received was accessioned into the Metabolon LIMS system and was assigned by the LIMS a unique identifier that was associated with the original source identifier only. This identifier was used to track all sample handling, tasks, results, etc. The samples (and all derived aliquots) were tracked by the LIMS system. All portions of any sample were automatically assigned their own unique identifiers by the LIMS when a new task was created; the relationship of these samples was also tracked. All samples were maintained at -80°C until processed.

#### Sample Preparation

Samples were prepared using the automated MicroLab STAR® system from Hamilton Company. Several recovery standards were added prior to the first step in the extraction process for QC purposes. To remove protein, dissociate small molecules bound to protein or trapped in the precipitated protein matrix, and to recover chemically diverse metabolites, proteins were precipitated with methanol under vigorous shaking for 2 min (Glen Mills GenoGrinder 2000) followed by centrifugation. The resulting extract was divided into five fractions: two for analysis by two separate reverse phase (RP)/UPLC-MS/MS methods with positive ion mode electrospray ionization (ESI), one for analysis by RP/UPLC-MS/MS with negative ion mode ESI, one for analysis by HILIC/UPLC-MS/MS with negative ion mode ESI, and one sample was reserved for backup. Samples were placed briefly on a TurboVap® (Zymark) to remove the organic solvent. The sample extracts were stored overnight under nitrogen before preparation for analysis.

#### QA/QC

Several types of controls were analyzed in concert with the experimental samples: a pooled matrix sample generated by taking a small volume of each experimental sample (or alternatively, use of a pool of well-characterized human plasma) served as a technical replicate throughout the data set; extracted water samples served as process blanks; and a cocktail of QC standards that were carefully chosen not to interfere with the measurement of endogenous compounds were spiked into every analyzed sample, allowed instrument performance monitoring and aided chromatographic alignment. Supplemental Methods Tables 1 and 2 describe these QC samples and standards. Instrument variability was determined by calculating the median relative standard deviation (RSD) for the standards that were added to each sample prior to injection into the mass spectrometers. Overall process variability was determined by calculating the median RSD for all endogenous metabolites (i.e., non-instrument standards) present in 100% of the pooled matrix samples. Experimental samples were randomized across the platform run with QC samples spaced evenly among the injections, as outlined in Supplemental Methods Figure 1.

**Supplemental Methods Table 1:** Description of Metabolon QC Samples.

| Type | Description | Purpose |
| --- | --- | --- |
| MTRX | Large pool of human plasma maintained by Metabolon that has been characterized extensively. | Assure that all aspects of the Metabolon process are operating within specifications. |
| CMTRX | Pool created by taking a small aliquot from every customer sample. | Assess the effect of a non-plasma matrix on the Metabolon process and distinguish biological variability from process variability. |
| PRCS | Aliquot of ultra-pure water | Process Blank used to assess the contribution to compound signals from the process. |
| SOLV | Aliquot of solvents used in extraction. | Solvent Blank used to segregate contamination sources in the extraction. |

**Supplemental Methods Table 2:** Metabolon QC Standards.

| Type | Description | Purpose |
| --- | --- | --- |
| RS | Recovery Standard | Assess variability and verify performance of extraction and instrumentation. |
| IS | Internal Standard | Assess variability and performance of instrument. |

**Supplemental Methods Figure 1.**
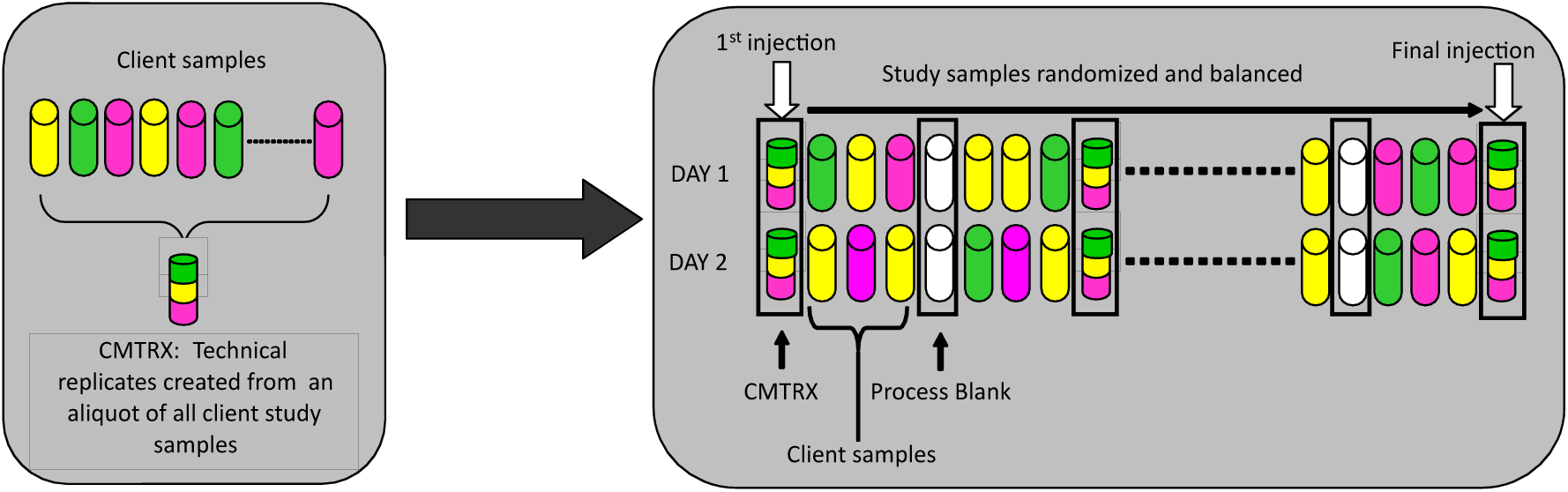
Preparation of client-specific technical replicates. A small aliquot of each client sample (colored cylinders) is pooled to create a CMTRX technical replicate sample (multi-colored cylinder), which is then injected periodically throughout the platform run. Variability among consistently detected biochemicals can be used to calculate an estimate of overall process and platform variability.

### Ultrahigh Performance Liquid Chromatography-Tandem Mass Spectroscopy (UPLC-MS/MS

All methods utilized a Waters ACQUITY ultra-performance liquid chromatography (UPLC) and a Thermo Scientific Q-Exactive high resolution/accurate mass spectrometer interfaced with a heated electrospray ionization (HESI-II) source and Orbitrap mass analyzer operated at 35,000 mass resolution. The sample extract was dried then reconstituted in solvents compatible to each of the four methods. Each reconstitution solvent contained a series of standards at fixed concentrations to ensure injection and chromatographic consistency. One aliquot was analyzed using acidic positive ion conditions, chromatographically optimized for more hydrophilic compounds. In this method, the extract was gradient eluted from a C18 column (Waters UPLC BEH C18-2.1x100 mm, 1.7 µm) using water and methanol, containing 0.05% perfluoropentanoic acid (PFPA) and 0.1% formic acid (FA). Another aliquot was also analyzed using acidic positive ion conditions, however it was chromatographically optimized for more hydrophobic compounds. In this method, the extract was gradient eluted from the same afore mentioned C18 column using methanol, acetonitrile, water, 0.05% PFPA and 0.01% FA and was operated at an overall higher organic content. Another aliquot was analyzed using basic negative ion optimized conditions using a separate dedicated C18 column. The basic extracts were gradient eluted from the column using methanol and water, however with 6.5mM Ammonium Bicarbonate at pH 8. The fourth aliquot was analyzed via negative ionization following elution from a HILIC column (Waters UPLC BEH Amide 2.1x150 mm, 1.7 µm) using a gradient consisting of water and acetonitrile with 10mM Ammonium Formate, pH 10.8. The MS analysis alternated between MS and data-dependent MS^n^ scans using dynamic exclusion. The scan range varied slighted between methods but covered 70-1000 m/z. Raw data files are archived and extracted as described below.

### Bioinformatics

The informatics system consisted of four major components, the Laboratory Information Management System (LIMS), the data extraction and peak-identification software, data processing tools for QC and compound identification, and a collection of information interpretation and visualization tools for use by data analysts. The hardware and software foundations for these informatics components were the LAN backbone, and a database server running Oracle 10.2.0.1 Enterprise Edition.

### LIMS

The purpose of the Metabolon LIMS system was to enable fully auditable laboratory automation through a secure, easy to use, and highly specialized system. The scope of the Metabolon LIMS system encompasses sample accessioning, sample preparation and instrumental analysis and reporting and advanced data analysis. All of the subsequent software systems are grounded in the LIMS data structures. It has been modified to leverage and interface with the in-house information extraction and data visualization systems, as well as third party instrumentation and data analysis software.

### Data Extraction and Compound Identification

Raw data was extracted, peak-identified and QC processed using Metabolon’s hardware and software. These systems are built on a web-service platform utilizing Microsoft’s .NET technologies, which run on high-performance application servers and fiber-channel storage arrays in clusters to provide active failover and load-balancing. Compounds were identified by comparison to library entries of purified standards or recurrent unknown entities. Metabolon maintains a library based on authenticated standards that contains the retention time/index (RI), mass to charge ratio (m/z), and chromatographic data (including MS/MS spectral data) on all molecules present in the library. Furthermore, biochemical identifications are based on three criteria: retention index within a narrow RI window of the proposed identification, accurate mass match to the library +/- 10 ppm, and the MS/MS forward and reverse scores between the experimental data and authentic standards. The MS/MS scores are based on a comparison of the ions present in the experimental spectrum to the ions present in the library spectrum. While there may be similarities between these molecules based on one of these factors, the use of all three data points can be utilized to distinguish and differentiate biochemicals. More than 3300 commercially available purified standard compounds have been acquired and registered into LIMS for analysis on all platforms for determination of their analytical characteristics. Additional mass spectral entries have been created for structurally unnamed biochemicals, which have been identified by virtue of their recurrent nature (both chromatographic and mass spectral). These compounds have the potential to be identified by future acquisition of a matching purified standard or by classical structural analysis.

### Curation

A variety of curation procedures were carried out to ensure that a high quality data set was made available for statistical analysis and data interpretation. The QC and curation processes were designed to ensure accurate and consistent identification of true chemical entities, and to remove those representing system artifacts, mis-assignments, and background noise. Metabolon data analysts use proprietary visualization and interpretation software to confirm the consistency of peak identification among the various samples. Library matches for each compound were checked for each sample and corrected if necessary.

### Metabolite Quantification and Data Normalization

Peaks were quantified using area-under-the-curve. For studies spanning multiple days, a data normalization step was performed to correct variation resulting from instrument inter-day tuning differences. Essentially, each compound was corrected in run-day blocks by registering the medians to equal one (1.00) and normalizing each data point proportionately (termed the “block correction”; Figure 2). For studies that did not require more than one day of analysis, no normalization is necessary, other than for purposes of data visualization. In certain instances, biochemical data may have been normalized to an additional factor (e.g., cell counts, total protein as determined by Bradford assay, osmolality, etc.) to account for differences in metabolite levels due to differences in the amount of material present in each sample.

**Supplemental Methods Figure 2:**
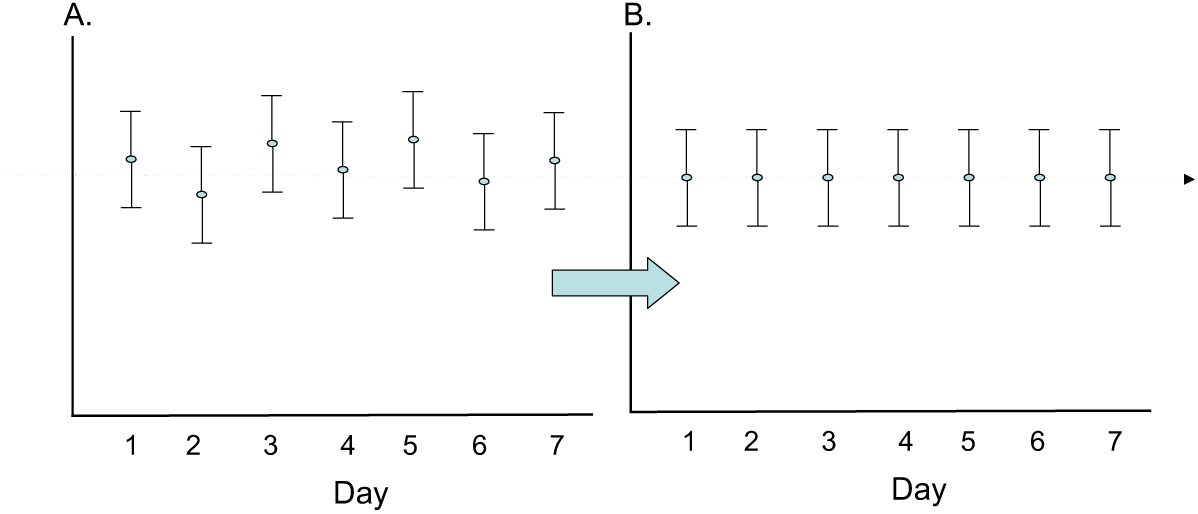
Visualization of data normalization steps for a multiday platform run.

### Statistical Methods and Terminology

#### Statistical Calculations

For many studies, two types of statistical analysis are usually performed: (1) significance tests and (2) classification analysis. Standard statistical analyses are performed in ArrayStudio on log transformed data. For those analyses not standard in ArrayStudio, the programs R (http://cran.r-project.org/) or JMP are used. Below are examples of frequently employed significance tests and classification methods followed by a discussion of p- and q-value significance thresholds.

### **1.** Welch’s two-sample t-test

Welch’s two-sample t-test is used to test whether two unknown means are different from two independent populations.

This version of the two-sample t-test allows for unequal variances (variance is the square of the standard deviation) and has an approximate t-distribution with degrees of freedom estimated using Satterthwaite’s approximation. The test statistic is given by 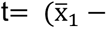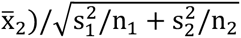, and the degrees of freedom is given by 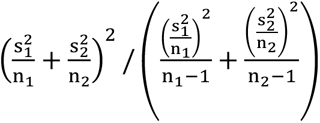 where x̄_1_, x̄_2_ are the sample means, s1, s2, are the sample standard deviations, and n1, n2 are the samples sizes from groups 1 and 2, respectively. We typically use a two-sided test (tests whether the means are different) as opposed to a one-sided test (tests whether one mean is greater than the other).

### **2.** p-values

For statistical significance testing, p-values are given. The lower the p-value, the more evidence we have that the null hypothesis (typically that two population means are equal) is not true. If “statistical significance” is declared for p-values less than 0.05, then 5% of the time we incorrectly conclude the means are different, when actually they are the same.

The p-value is the probability that the test statistic is at least as extreme as observed in this experiment given that the null hypothesis is true. Hence, the more extreme the statistic, the lower the p-value and the more evidence the data gives against the null hypothesis.

### **3.** q-values

The level of 0.05 is the false positive rate when there is one test. However, for a large number of tests we need to account for false positives. There are different methods to correct for multiple testing. The oldest methods are family-wise error rate adjustments (Bonferroni, Tukey, etc.), but these tend to be extremely conservative for a very large number of tests. With gene arrays, using the False Discovery Rate (FDR) is more common. The family-wise error rate adjustments give one a high degree of confidence that there are zero false discoveries. However, with FDR methods, one can allow for a small number of false discoveries. The FDR for a given set of compounds can be estimated using the q-value (see Storey J and Tibshirani R. (2003) Statistical significance for genomewide studies. Proc. Natl. Acad. Sci. USA 100: 9440-9445; PMID: 12883005).

In order to interpret the q-value, the data must first be sorted by the p-value then choose the cutoff for significance (typically p<0.05). The q-value gives the false discovery rate for the selected list (i.e., an estimate of the proportion of false discoveries for the list of compounds whose p-value is below the cutoff for significance). For Supplemental Methods Table 3 below, if the whole list is declared significant, then the false discovery rate is approximately 10%. If everything from Compound 079 and above is declared significant, then the false discovery rate is approximately 2.5%.

**Supplemental Methods Table 3:**
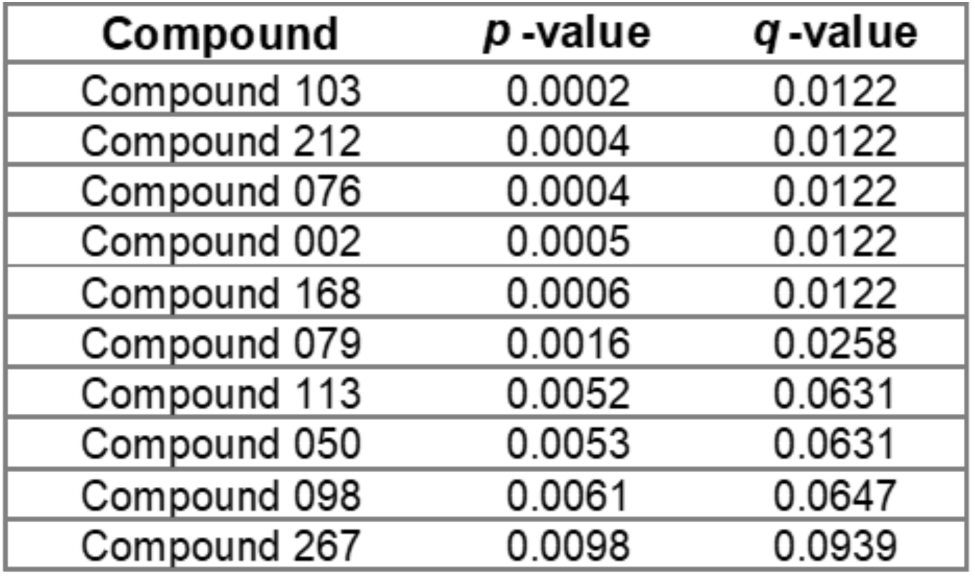
Example of q-value interpretation.

| Compound | *p* -value | *q* -value |
| --- | --- | --- |
| Compound 103 | 0.0002 | 0.0122 |
| Compound 212 | 0.0004 | 0.0122 |
| Compound 076 | 0.0004 | 0.0122 |
| Compound 002 | 0.0005 | 0.0122 |
| Compound 168 | 0.0006 | 0.0122 |
| Compound 079 | 0.0016 | 0.0258 |
| Compound 113 | 0.0052 | 0.0631 |
| Compound 050 | 0.0053 | 0.0631 |
| Compound 098 | 0.0061 | 0.0647 |
| Compound 267 | 0.0098 | 0.0939 |

### **4.** Random Forest

Random forest is a supervised classification technique based on an ensemble of decision trees (see Breiman L. (2001) Random Forests. Machine Learning. 45: 5-32; http://link.springer.com/article/10.1023%2FA%3A1010933404324). For a given decision tree, a random subset of the data with identifying true class information is selected to build the tree (“bootstrap sample” or “training set”), and then the remaining data, the “out-of-bag” (OOB) variables, are passed down the tree to obtain a class prediction for each sample. This process is repeated thousands of times to produce the forest. The final classification of each sample is determined by computing the class prediction frequency (“votes”) for the OOB variables over the whole forest. For example, suppose the random forest consists of 50,000 trees and that 25,000 trees had a prediction for sample 1. Of these 25,000, suppose 15,000 trees classified the sample as belonging to Group A and the remaining 10,000 classified it as belonging to Group B. Then the votes are 0.6 for Group A and 0.4 for Group B, and hence the final classification is Group A. This method is unbiased since the prediction for each sample is based on trees built from a subset of samples that do not include that sample. When the full forest is grown, the class predictions are compared to the true classes, generating the “OOB error rate” as a measure of prediction accuracy. Thus, the prediction accuracy is an unbiased estimate of how well one can predict sample class in a new data set. Random forest has several advantages – it makes no parametric assumptions, variable selection is not needed, it does not overfit, it is invariant to transformation, and it is fairly easy to implement with R.

To determine which variables (biochemicals) make the largest contribution to the classification, a “variable importance” measure is computed. We use the “Mean Decrease Accuracy” (MDA) as this metric. The MDA is determined by randomly permuting a variable, running the observed values through the trees, and then reassessing the prediction accuracy. If a variable is not important, then this procedure will have little change in the accuracy of the class prediction (permuting random noise will give random noise). By contrast, if a variable is important to the classification, the prediction accuracy will drop after such a permutation, which we record as the MDA. Thus, the random forest analysis provides an “importance” rank ordering of biochemicals; we typically output the top 30 biochemicals in the list as potentially worthy of further investigation.

**S2 Supplemental Table.**
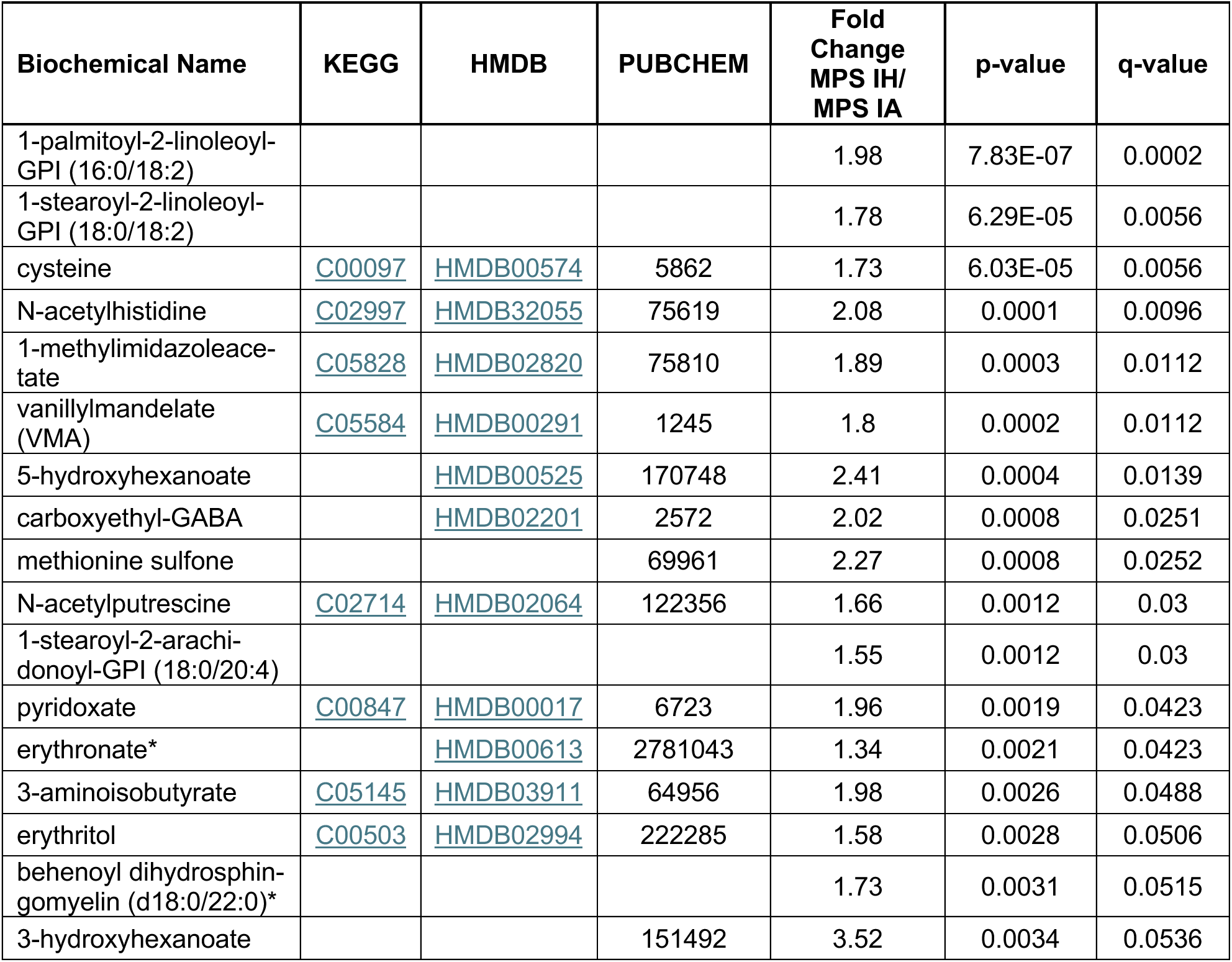
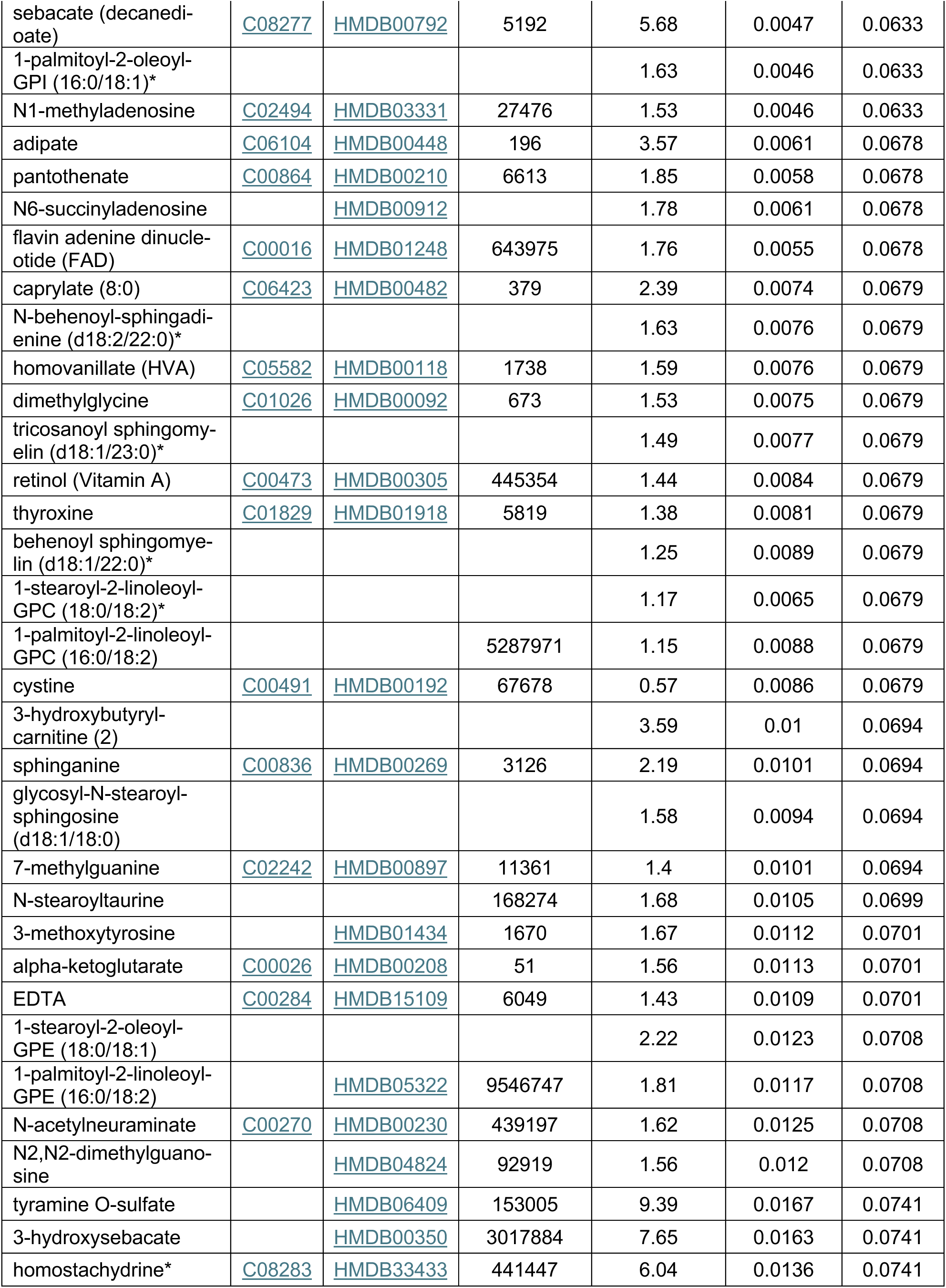
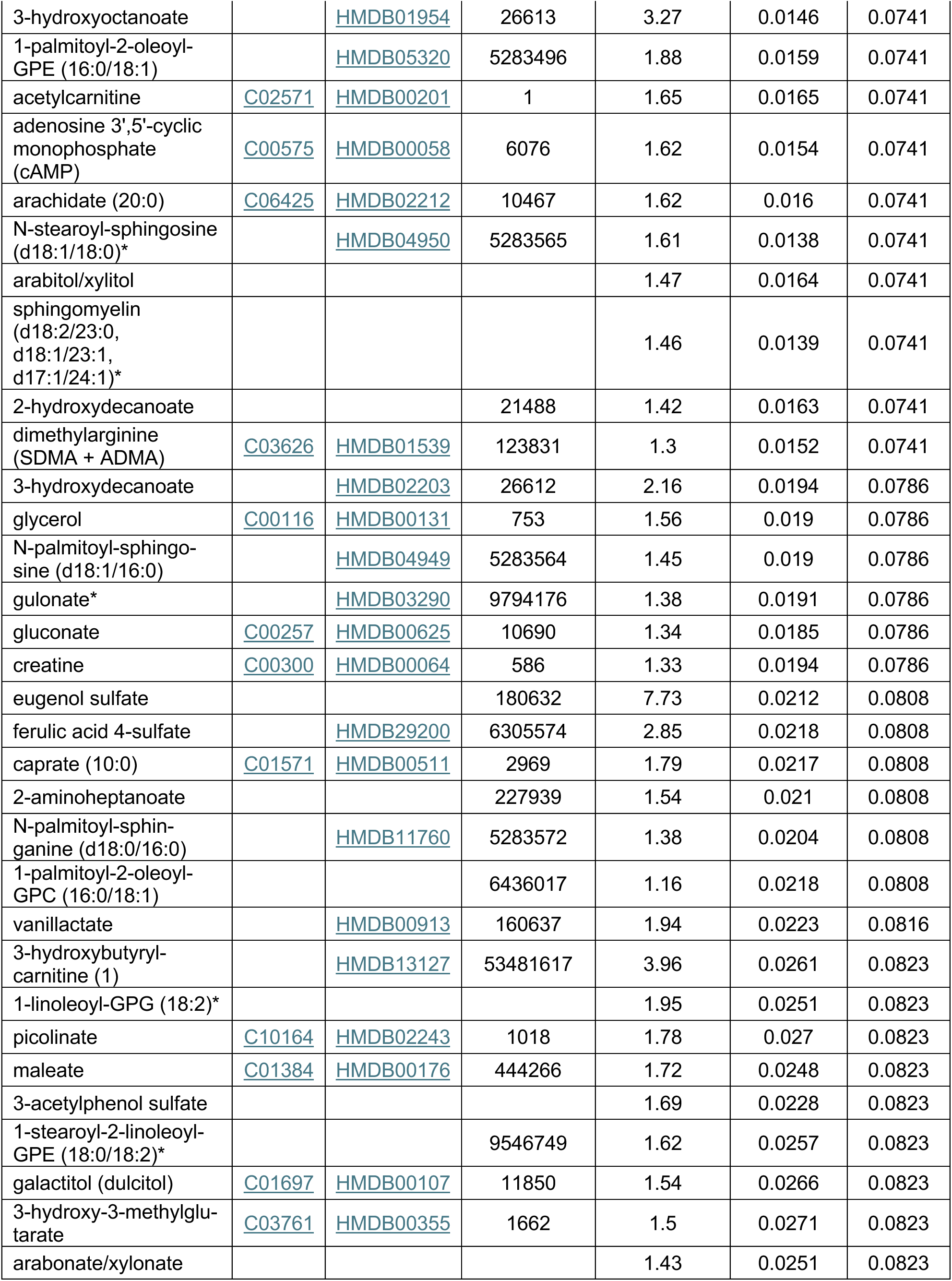
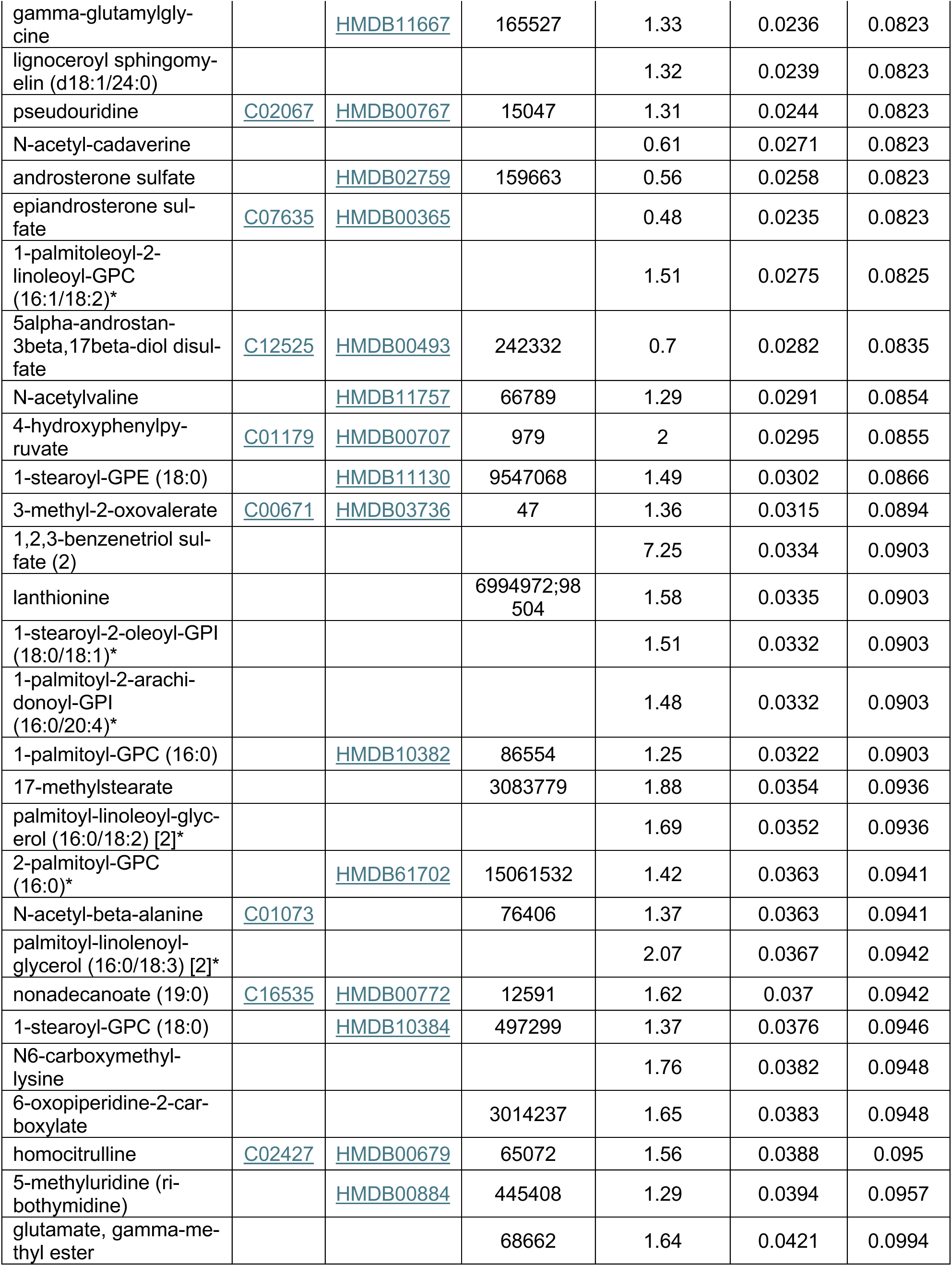
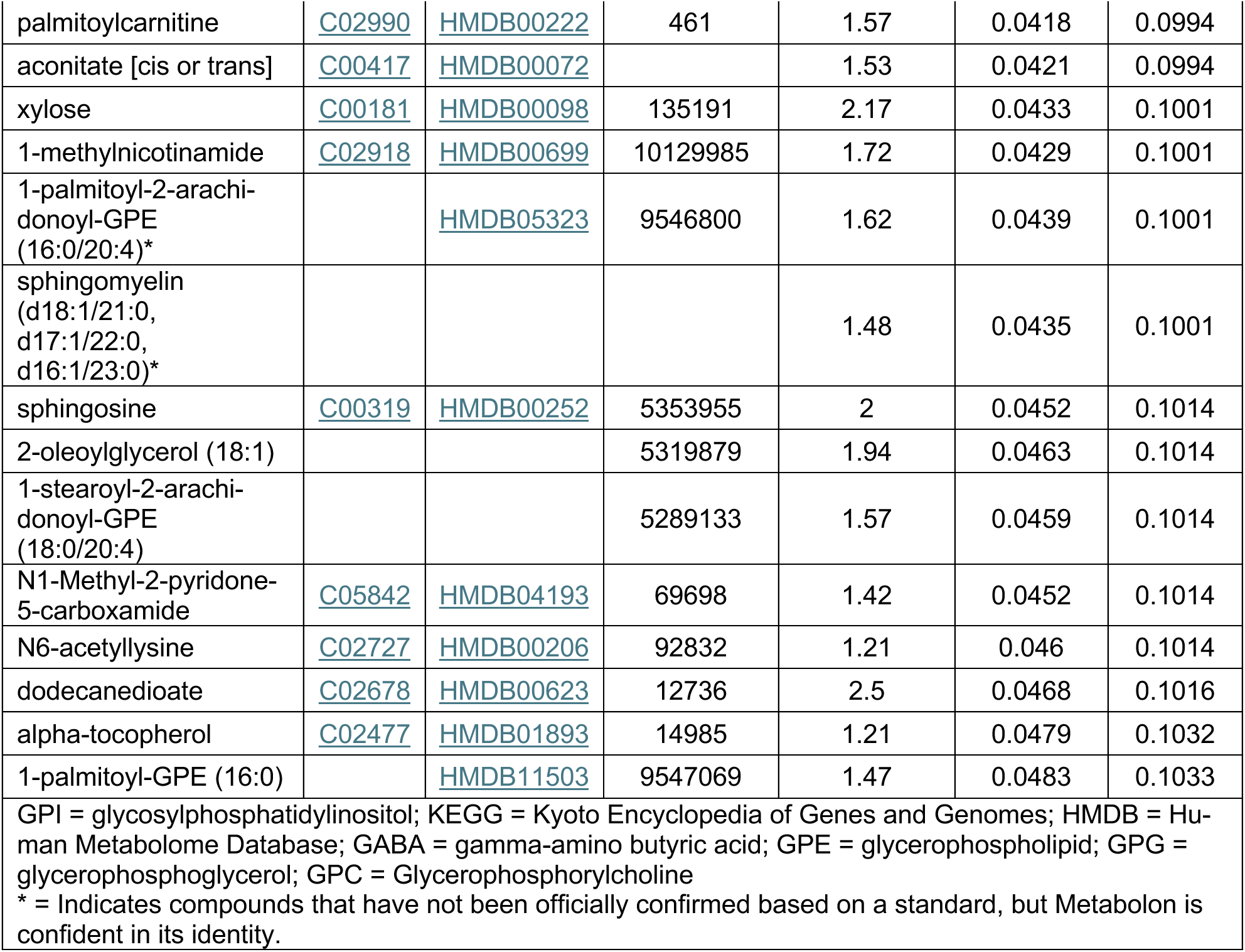
Metabolites with a p-value < 0.05 in comparative metabolomic analysis.

